# Novel delivery of cellular therapy to reduce ischaemia reperfusion injury in kidney transplantation

**DOI:** 10.1101/19005546

**Authors:** Emily R Thompson, Lucy Bates, Ibrahim K Ibrahim, Avinash Sewpaul, Ben Stenberg, Andrew McNeill, Rodrigo Figueiredo, Tom Girdlestone, Georgina C Wilkins, Ellen A Irwin, Samuel J Tingle, William E Scott, Henrique Lamos, Andrew L. Mellor, Valerie D. Roobrouck, Anthony E. Ting, Sarah A Hosgood, Michael L Nicholson, Andrew J Fisher, Simi Ali, Neil S Sheerin, Colin H Wilson

**Author notes:** Institute of Cellular Medicine Newcastle University, UK, NE2 4HH.

## Abstract

*Ex-vivo* normothermic machine perfusion (NMP) of donor kidneys prior to transplantation provides a platform for direct delivery of cellular therapeutics to optimise organ quality prior to transplantation. Multipotent Adult Progenitor Cells (MAPC^®^) possess potent immunomodulatory properties which could prove beneficial in minimising subsequent ischaemia reperfusion injury. We investigated the potential reconditioning capability of MAPC cells in kidney NMP.

**Methods:** Pairs (5) of human kidneys from the same donor were simultaneously perfused for 7 hours. The right or left kidney was randomly allocated to receive MAPC treatment. Serial samples of perfusate, urine and tissue biopsies were taken for comparison with the control paired kidney.

**Results:** MAPC-treated kidneys demonstrated improved urine output (p<0.01), decreased expression of the kidney injury biomarker NGAL (p<0.01), improved microvascular perfusion on contrast enhanced ultrasound (cortex p<0.05, medulla p<0.01), downregulation of IL-1β (p<0.05) and upregulation of IL-10 (p<0.05) and Indolamine-2, 3-dioxygenase (p<0.05). A mouse model of intraperitoneal chemotaxis demonstrated decreased neutrophil recruitment when stimulated with perfusate from MAPC-treated kidneys (p<0.01). Immunofluorescence revealed pre-labelled MAPC cells home to the perivascular space in the kidneys during NMP. MAPC therapy was not associated with detrimental physiological or embolic events.

**Conclusion:** We report the first successful delivery of cellular therapy to a kidney during NMP. Kidneys treated with MAPC cells demonstrate improvement in clinically relevant functional parameters and injury biomarkers. This novel method of cell therapy delivery provides an exciting opportunity to recondition organs prior to clinical transplantation.

**One Sentence Summary:** Ex-vivo reconditioning of human kidneys using Multipotent Adult Progenitor Cell therapy delivered during normothermic machine perfusion.

## Introduction

Currently the UK kidney transplant waiting list stands at over 5000 patients and the average waiting time is 3 years^1^. The organ shortage has led to increased use of organs from donation after circulatory death (DCD) and extended criteria donors (ECD) to bridge the gap between supply and demand ^2^. Concerns regarding inferior transplant outcomes from DCD & ECD organs can lead to under utilization of this valuable resource ^3, 4^. DCD & ECD organs are more susceptible to damage from ischaemia reperfusion injury (IRI) manifesting as delayed graft function (DGF) and this can diminish graft survival ^5, 6^. IRI is the result of hypoxia followed by restoration of blood flow resulting in microvascular dysfunction, inflammation, immune activation and tissue injury ^7^. IRI is an unavoidable consequence of solid organ transplantation and results in DGF in up to 50% of kidney transplants ^8^. As the transplant community becomes increasing reliant on marginal donors, new therapeutic approaches to reduce IRI and optimise utilisation of ECD kidneys are leading to a greater focus on improving organ preservation.

Normothermic Machine Perfusion (NMP) is a method of organ preservation that facilitates restoration of cellular metabolism, reviving the organ *ex vivo* to resume normal physiological functions ^9^. This method has many potential benefits over traditional, static cold storage (SCS), including the opportunity to undertake real-time objective assessments of organ quality prior to transplantation. Over the past 10 years, a number of NMP techniques and commercially available devices have been adopted into clinical practice for kidney, heart, liver and lung transplantation ^10, 11^. Nasralla et al recently reported the first international, multicentre, randomised controlled trial of 220 liver transplants investigating liver NMP for organ preservation. This study clearly demonstrated NMP had significant advantages compared with SCS. There was a 50% reduction in graft injury (measured by hepatocellular enzyme release), a 50% reduction in organ discard, and a safe increase in preservation times ^12^. This promising study has confirmed NMP as a viable, realistic technology with wider implications for its translation into other solid organ types and organ reconditioning.

Kidney NMP was first described in 2008 ^13^. In this system, a paediatric cardiopulmonary bypass machine and membrane oxygenator provides an *ex vivo* kidney with oxygenated red blood cells suspended in crystalloid at 37°C ^14^. A national multicentre phase 3 randomised controlled trial is currently underway to investigate the effectiveness of this technology in reducing DGF rates^15^.

NMP provides a unique opportunity to deliver organ-directed, reconditioning therapies. By establishing an isolated *ex vivo* platform with a metabolically active organ, therapies targeting ischaemia reperfusion injury can be delivered to the kidney ^16^. This could prove to be transformative, allowing delivery of stem cell or gene therapies directly to the kidney reducing off-target, systemic effects in recipients.

Multipotent Adult Progenitor Cells (MAPC) are adult, bone-marrow derived stromal cells first described in 2002 ^17^. MAPC cells represent an attractive ‘off-the-shelf’ cell therapy option as they lack MHC Class II, or co-stimulatory molecules (CD80, CD86 & CD40) and have low levels of MHC Class I expression meaning they are non-immunogenic ^18^. MAPC cells release anti-inflammatory, immunomodulatory and pro-tolerogenic cytokines thereby limiting infiltrating pathogenic immune cells ^19^. MAPC cells have also demonstrated potent *in vitro* immunosuppressive effects on T-cell proliferation ^20^. Maximal immunosuppressive effects were achieved in the most pro-inflammatory environments suggesting possible benefit in the highly inflammatory, marginal organ.

These *in vitro* findings have been validated by a number of animal models confirming the immunomodulatory capacity of MAPC cells in a transplant setting. In a rat allogeneic heterotopic heart transplant model (Lewis-into ACI) allogeneic MAPC cells could successfully replace standard pharmacological immunosuppression to maintain long-term graft survival mediated by local heart-specific effects of T regulatory cells ^21^.

There have also been a number of successful clinical trials harnessing the immunomodulatory potential of MAPC treatment for Graft versus Host Disease ^22^, Acute Respiratory Distress Syndrome ^23^, Myocardial Infarction ^24^ and Ischaemic Stroke ^25^. The studies in ischaemic stroke are currently the most advanced and there is an ongoing phase III study. In 2015, the first successful use of allogeneic MAPC therapy in human liver transplantation was reported ^26^. Interestingly, the recipient’s leucocyte population was profiled and revealed an increase in regulatory T cells on day 4 post-transplant. Associated with this was a downregulation of MHC Class II expression by CD14+ monocytes possibly representing diminished immune activation.

A number of animal models have explored the possibility of delivering a cell therapy during *ex vivo* NMP. A porcine model of lung NMP compared MAPC cells delivered intrabronchially to the left lung and demonstrated a reduction in pro-inflammatory cytokines and neutrophils in the bronchoalveolar lavage fluid after 6 hours of treatment when compared with the untreated right lung^27^. To date there have been no reported studies successfully administering a cell therapy to a human organ during *ex vivo* perfusion.

Our study aims to investigate the possible benefit of delivering MAPC cell therapy directly to marginal human kidneys in a pre-clinical NMP model.

## Results

### Kidneys included in pre-clinical series

5 pairs of kidneys (n=10) were included in the study (Table 1). The donors included represent a heterogeneous cohort with ages ranging from 52 to 77 years, but all were from either DCD donors or had characteristics consistent with ECD status. Cold ischaemic times were significantly extended due to the delays inherent in a kidney being offered for research only purposes. All the pairs of kidneys were declined for transplant due to a suspicion of non-renal malignancy identified at the retrieval operation.

**Table 1:** Clinical characteristics of donor kidney pairs included in study. DBD – Donation after Brainstem Death, DCD – Donation after Circulatory Death, CIT – Cold Ischaemic Time, L-left kidney, R – Right kidneys, A – artery, V – Vein, QAS – Quality Assessment Score (composite measure of kidney quality comprised of renal blood flow, urine output and macroscopic perfusion appearance. Scored 1-5(best = 1, worst = 5, suitable for transplant if score ≤3)

| Kidney pair | Age (years) | Donor Type | Reason for decline | CIT (time) | Vascular Anatomy | Weight (g) | QAS score @1hr |
| --- | --- | --- | --- | --- | --- | --- | --- |
| Pair 1 | 60 | DBD | non-renal malignancy | 20h 35m | L – 1A 1V | L - 270g | L - 3 |
|  |  |  |  |  | R – 1A 2V | R - 260g | R - 3 |
| Pair 2 | 52 | DBD | non-renal malignancy | 29h 35m | L – 1A 1V | L - 235g | L - 2 |
|  |  |  |  |  | R – 2A 1V | R - 220g | R - 2 |
| Pair 3 | 62 | DCD | non-renal malignancy | 36h 20m | L – 2A 1V | L - 285g | L - 5 |
|  |  |  |  |  | R – 1A 1V | R - 288g | R - 5 |
| Pair 4 | 76 | DBD | non-renal malignancy | 35h 38m | L – 1A 1V | L - 199g | L - 1 |
|  |  |  |  |  | R – 1A 1V | R - 235g | R - 1 |
| Pair 5 | 77 | DCD | non-renal malignancy | 13h 06m | L – 1A 1V | L - 331g | L - 1 |
|  |  |  |  |  | R – 1A 1V | R - 249g | R - 1 |

### Establishing feasibility of cell therapy delivery during kidney NMP

#### Renal Physiology

Serial measurements of physiological parameters were recorded during NMP. Perfusate samples were analysed in real-time to assess adequate oxygenation, metabolic requirements and biochemical parameters. These were compared between the pairs of kidneys, control vs MAPC treated, and demonstrated equivalent organ physiology associated with MAPC cell infusion. For clinically relevant markers, potassium, lactate, renal blood flow & renal resistance, the kidneys were well matched throughout the 7-hour perfusion timeline (Figure 1 Panel A-D).

**Figure 1:**
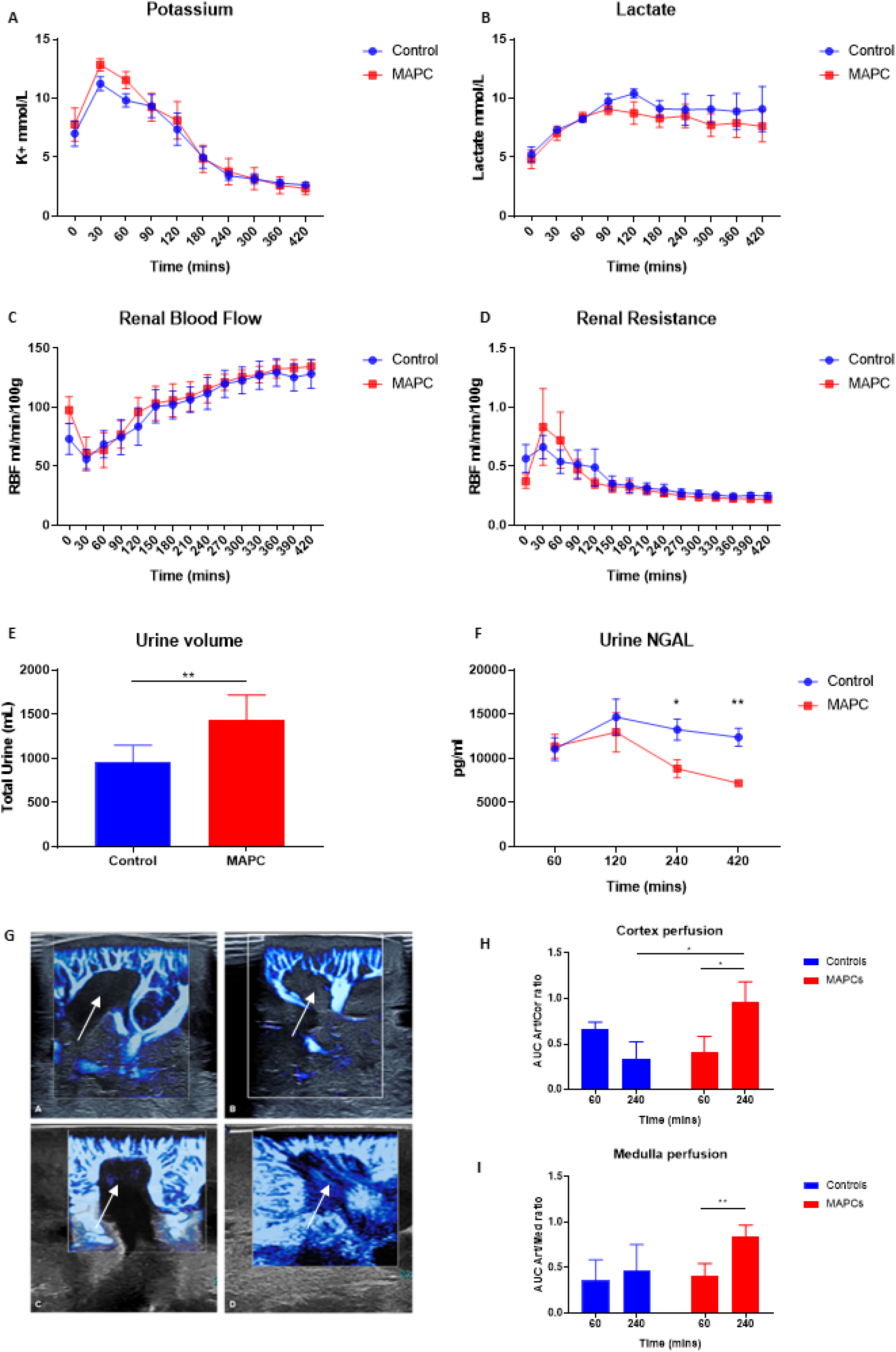
Establishing the feasibility of MAPC therapy during kidney NMP. Panels A-D depict real-time physiological parameters measured during the NMP timeline. Panel (A) electrolyte balance (potassium), (B) cellular metabolism (lactate), (C) renal blood flow and (D) intrarenal resistance recorded throughout the perfusion timeline. Panel E depicts the impact of MAPC treatment on urine output; n=5pairs, paired t-test, **p<0.01 (MAPC treated kidneys mean urine output 1436mL±281 vs control 960mL±189). Panel F demonstrates MAPC treatment effect on a biomarker of kidney injury NGAL concentration measured at serial time points during perfusion, repeated-measures two-way ANOVA with Sidak’s multiple comparison’s post-hoc test, *p<0.05 **p<0.01 (MAPC treated mean NGAL concentration 7196pg/ml ± 379 vs control 12400pg/ml ± 1014 at 7 hours). Panel G demonstrates still images taken from one pair of kidneys recorded during Doppler Ultrasound MicroFlow Imaging. Blue colouring represents aoverlaid Doppler signal from blood flow within microvasculature. Image A – Control kidney after 60 minutes of NMP, Image B – the same control kidney scanned after 4 hours of NMP. Image C MAPC treated kidney after 60 minutes of NMP before MAPC cell infusion, Image D-the same kidney scanned 4 hours after MAPC cell infusion demonstrating new blood flow within the medullary pyramid (white arrow). CEUS performed during NMP to assess the impact of MAPC cell therapy on microvascular perfusion in ischaemic kidneys. Panel H depicts objective AUC analysis of cortex perfusion comparing Contrast Enhanced Ultrasound scans performed at 60 minutes and 4 hours. Panel I depicts AUC analysis of medulla perfusion comparing CEUS scans performed at 60 minutes and 4 hours. n=3, paired t-test, *p<0.05 **<0.01 (MAPC treated mean cortex perfusion AUC 0.97±0.13 vs control 0.34±0.11).

During NMP the ureter of the kidney was cannulated and attached to a urometer to facilitate hourly urine output measurement and sampling. In the kidneys treated with MAPC cells there was significantly higher urine output compared to control kidneys during NMP, p<0.01 (Figure 1, Panel E).

#### Kidney Injury Biomarkers

There are a number of validated biomarkers regularly used in kidney transplant or acute kidney injury research, including Kidney Injury Marker 1 (KIM-1) and Neutrophil Gelatinase-Associated Lipocalin (NGAL) ^28^. With the advent of NMP technologies we can analyse these biomarkers during perfusion to evaluate reconditioning. In this study, we assessed both the levels of urinary KIM-1 and NGAL over time and compared control kidneys with the paired MAPC treated kidneys. This demonstrated that the MAPC treated kidneys showed a significantly lower urinary concentration of NGAL over the course of 7 hours NMP, p<0.01 (Figure 1, Panel F). Concentrations of urinary KIM-1 were matched between both treatment groups throughout perfusion (supplementary figure 1, Panel A).

#### Ex vivo Ultrasound

To determine if the administration of a MAPC cell bolus was associated with any effects on the renal microvasculature we performed a number of imaging studies using ultrasound. MicroFlow Imaging (MFI) Doppler ultrasound was performed. MFI is an ultrasound technology that can provide high resolution detail on blood flow within small vessels. This technique was used to assess blood flow following MAPC treatment and revealed restored blood flow within the renal medulla 4 hours after MAPC cell infusion (Figure 1, Panel G, indicated by white arrow). The same effect was not seen in control kidneys.

Alongside MFI, contrast enhanced ultrasound (CEUS) was performed after 60 minutes of NMP (before MAPC cell infusion) and 4 hours later. This was primarily to investigate if the cell bolus was associated with micro-emboli or occlusion of the renal microvasculature as the mean diameter of a MAPC cell is greater than a capillary. CEUS provides a quantifiable assessment of microvascular perfusion and, demonstrated a significant improvement in both cortical and medullary perfusion after 4 hours of MAPC treatment during NMP, compared to the control kidneys, p<0.05 (Figure 1 Panel H & I).

### Evaluating the immunomodulatory potential of MAPC therapy during NMP

#### Cytokine Profiling

MAPC cells are reported to mediate their immunomodulatory capacity through increasing anti-inflammatory cytokines and down regulating pro-inflammatory cytokines. A panel of cytokines were measured at serial time points during perfusion (time-zero, 1hr, 2hr, 4hr & 7hrs) to evaluate this phenomenon during NMP. This panel revealed MAPC treated kidneys had a significant reduction in IL-1β, p<0.05 (Figure 2 Panel A). There was also a significant increase in anti-inflammatory cytokine IL-10, p<0.05 (Figure 2, Panel B). There was also a non-significant reduction in other pro-inflammatory cytokines; IL-6, IL-1α, IL-17 & IL-8 but his was not the case for all cytokines; TNFα, MIP-1β, IL-2, IFNγ. (Supplementary Figure 1, Panel B)

**Figure 2:**
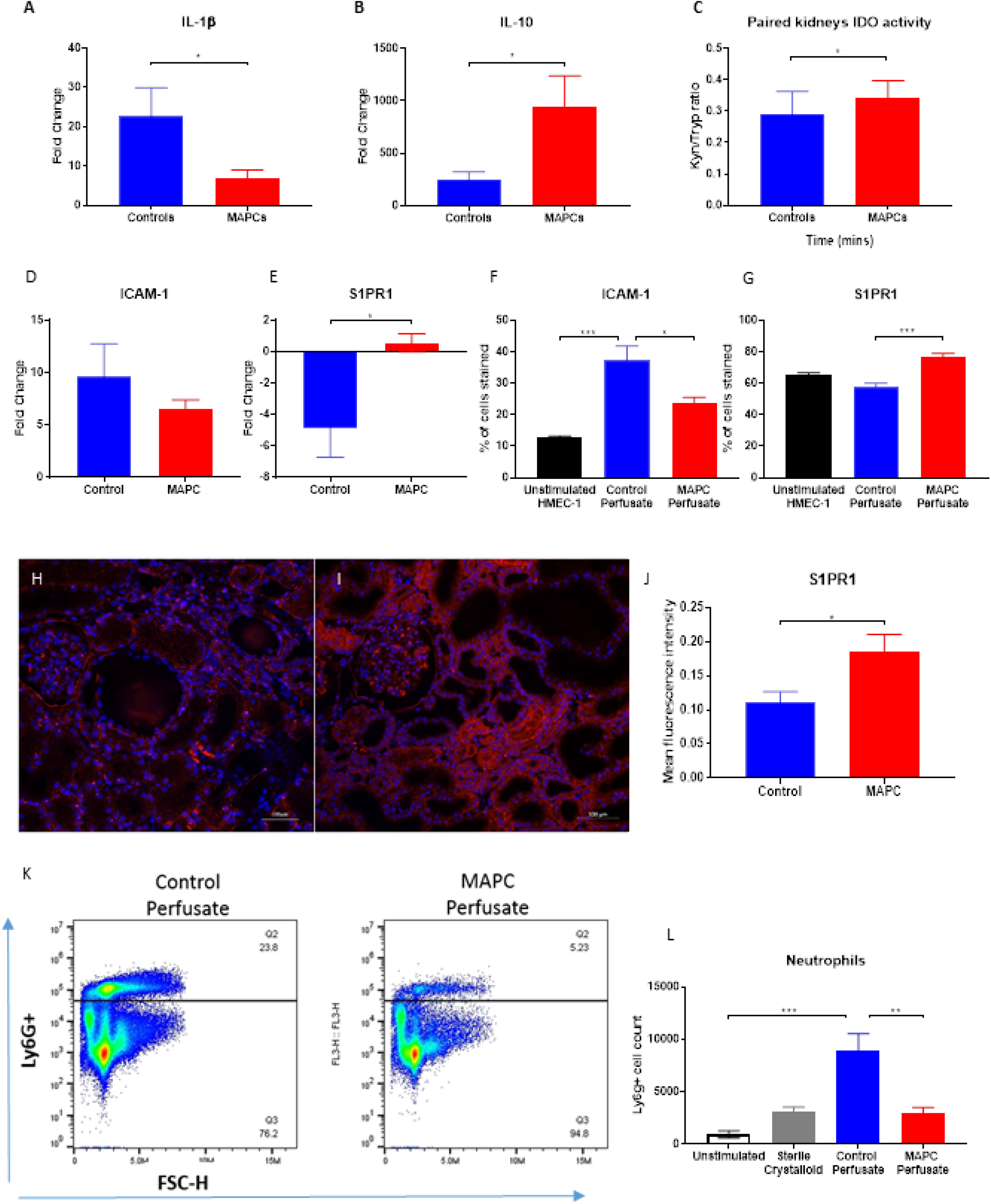
Evaluating the immunomodulatory capability of MAPC therapy during kidney NMP. Panels A & B depict significant results from custom Mesoscale Discovery™ panel analyzing perfusate cytokine concentrations comparing control (blue) with MAPC treated (red) NMP kidneys. Results are expressed as the mean fold change calculated relative to time zero from each kidney. Panel A demonstrates a significant decreased expression of cytokine IL-1β, paired t-test, n=5pairs, *p<0.05 (MAPC treated mean fold change 6.9±2.1 vs control 26.8±7.7). Panel B demonstrates a significant increase cytokine IL-10 in the MAPC treated group, paired t-test, n=5 pairs, *p<0.05 (MAPC treated mean fold change 949.4±288.6 vs control 252.9±71.3). Panel C compares IDO activity in paired kidneys after 7 hours of NMP, paired t-test, n=5 pairs, *p<0.05 (MAPC treated K/T ratio 0.34±0.02 vs control 0.29±0.03). Panel D & E depict qPCR results from stimulated HMEC-1 cells treated with aceullular perfusate taken after 7 hours NMP from kidney pairs (control kidneys vs MAPC treated). Gene expression was calculated in reference to housekeeping gene GAPDH and unstimulated HMEC-1 cells grown to confluence. Panel D demonstrates the impact of MAPC perfusate secretome on HMEC-1 cell ICAM-1 gene expression, paired t-test, n=5 pairs, p=ns (MAPC treated mean fold change on qPCR 9.6±3.1 vs 6.5±0.9). Panel E demonstrates the impact of MAPC treated perfusate on HMEC-1 cell S1PR1 gene expression, paired t-test, *p<0.05 (MAPC treated mean fold change on qPCR −4.9±1.7 vs control 0.6±0.6). Panel F & G depict HMEC-1 cells immunohistochemistry staining for ICAM-1 & SIPR1 protein expression after stimulation with acellular perfusates and the percentage of positive staining cells (control vs MAPC treated). Graph F demonstrates comparison of ICAM-1 protein expression between treatment groups (MAPC treated 23.9±3.3 vs control perfusate 37.3±9.2). Graph G demonstrates comparison of S1PR1 protein expression between treatment groups (MAPC treated 76.5±4.7 vs control perfusate 57.6±4.9). Analysed with one-way ANOVA and Tukey’s test post-hoc analysis, *p<0.05, **p<0.01, ***p<0.001. Panel H & I depict S1PR1 staining visualized using fluorescence microscopy on kidney biopsies taken during NMP, blue stain is DAPI, red is S1PR1. Panel J demonstrates a significant upregulation of S1PR1 protein expression as quantified by mean fluorescence intensity, paired t-test, n=5 pairs, *p<0.05 (MAPC treated 0.11±0.01 vs control 0.18±0.02). Panels K&L demonstrate the results from the mouse intraperitoneal chemotaxis experiments. Panel K depicts typical flow cytometry scatter plots examining staining for Ly6G+ neutrophils isolated from peritoneal lavage for the different treatment groups. Panel L demonstrates the mean number of Ly6G+ cells isolated from the lavage samples. There is a significant reduction in intraperitoneal neutrophil recruitment in those mice stimulated with MAPC perfusate when compared to control perfusate, paired t-test, n=5 pairs, **p<0.01 (MAPC 2990±506 neutrophils vs control mice 8849±1675 neutrophils).

#### Indolamine-2, 3-dioxygenase activity

MAPC cells *in vitro* and *in vivo* are reported to regulate T cells via indolamine-2,3-dioxygenase (IDO) (19). Cells expressing IDO catabolize tryptophan (Trp) to suppress effector T cells and activate Foxp3-lineage regulatory CD4 T cells (Tregs) ^29^. IDO activity was assessed by measuring Trp and the catabolite kynurenine (Kyn) in the perfusate using HPLC. Data revealed that MAPC treated kidneys had significantly higher IDO activity (elevated Kyn:Trp ratio) following 7 hours of perfusion when compared to IDO activity in paired control kidneys, p<0.05 (Figure 2, Panel C).

#### MAPC perfusate secretome effect on HMEC-1 endothelial cell line model

An *in vitro* endothelial cell line model was used to better understand the impact of the MAPC secretome on kidney vascular endothelium and microvascular integrity. As MAPC cells are a biologically reactive product, the secretome produced by the cells is dependent upon the microenvironment in which they are delivered. Indeed, this is one of the attractive mechanisms of action we hope to harness through MAPC cell delivery to a marginal kidney during NMP - the cells should react to this pro-inflammatory, ischaemic environment and produce the required growth factors and mediators for optimum reconditioning. To model this *in vitro*, samples of acellular perfusate taken after 7 hours of NMP were added to Human Microvascular Endothelial Cells (HMEC-1) cells in culture. ICAM-1 (activation status) and S1PR1 (microvascular barrier integrity) expression was analysed in the HMEC-1 cells in response to perfusate from pairs of kidneys (untreated vs MAPC treated) (Figure 2 Panel D-G). ICAM-1 protein expression was significantly increased in the control group, p<0.001. However, in the MAPC treated group this upregulation was not as marked, p<0.05. In contrast, S1PR1 gene and protein expression in HMEC-1 cells was downregulated by control perfusate, however, MAPC perfusate maintained S1PR1 gene expression at unstimulated levels, p<0.05; and increased protein expression when compared with control perfusate, p<0.001). This preservation of S1PR1 protein was also seen on immunofluorescence in the tissue of NMP perfused kidneys, p<0.05 (Figure 2, Panel H-J).

#### MAPC secretome effect on mouse intraperitoneal chemotaxis assay

To evaluate if the MAPC secretome in the perfusate during NMP had an impact on leucocyte chemotaxis we used a small animal model. Samples of acellular perfusate were injected intraperitoneally to mice. There were 4 treatment groups, each containing 5 mice, as described in the methods section. Six hours after injection the intraperitoneal space was lavaged to harvest the immune cell infiltrate which was analysed using flow cytometry (supplementary figure 1, Panel C-E). CD45+ was used as a leucocyte marker and Ly6G+ for neutrophils. Control perfusate led to a significant increase in peritoneal neutrophils compared to crystalloid injections alone. This increase was not seen in perfusate from MAPC treated kidneys, p<0.01 (Figure 2, Panel J-L).

### Determining the physical distribution of NMP administered MAPC cells

MAPC cells were pre-labelled with a red fluorescent dye in order to understand cell fate following intra-arterial delivery during kidney NMP. To achieve this, following 7 hours of perfusion, the kidneys were de-cannulated and anatomical regions of interest sampled. Samples were taken from the cortex, medulla, artery and collecting system of the kidney and visualised using fluorescent immunohistochemistry and confocal microscopy. Nuclear counter-staining was with DAPI (blue). Additional staining for endothelial cell marker (CD31 - green) and a proximal tubular epithelial cell marker (Aquaporin-1) was performed to aid co-localisation of MAPC cells within the kidney’s architecture. This revealed that the majority of the red labelled MAPC cells were to be found in the glomeruli and around the peritubular capillaries kidneys, (Figure 3, Panel A-D). MAPC cells were also seen in the perivascular interstitial space, (Figure 3, Panel E).

**Figure 3:**
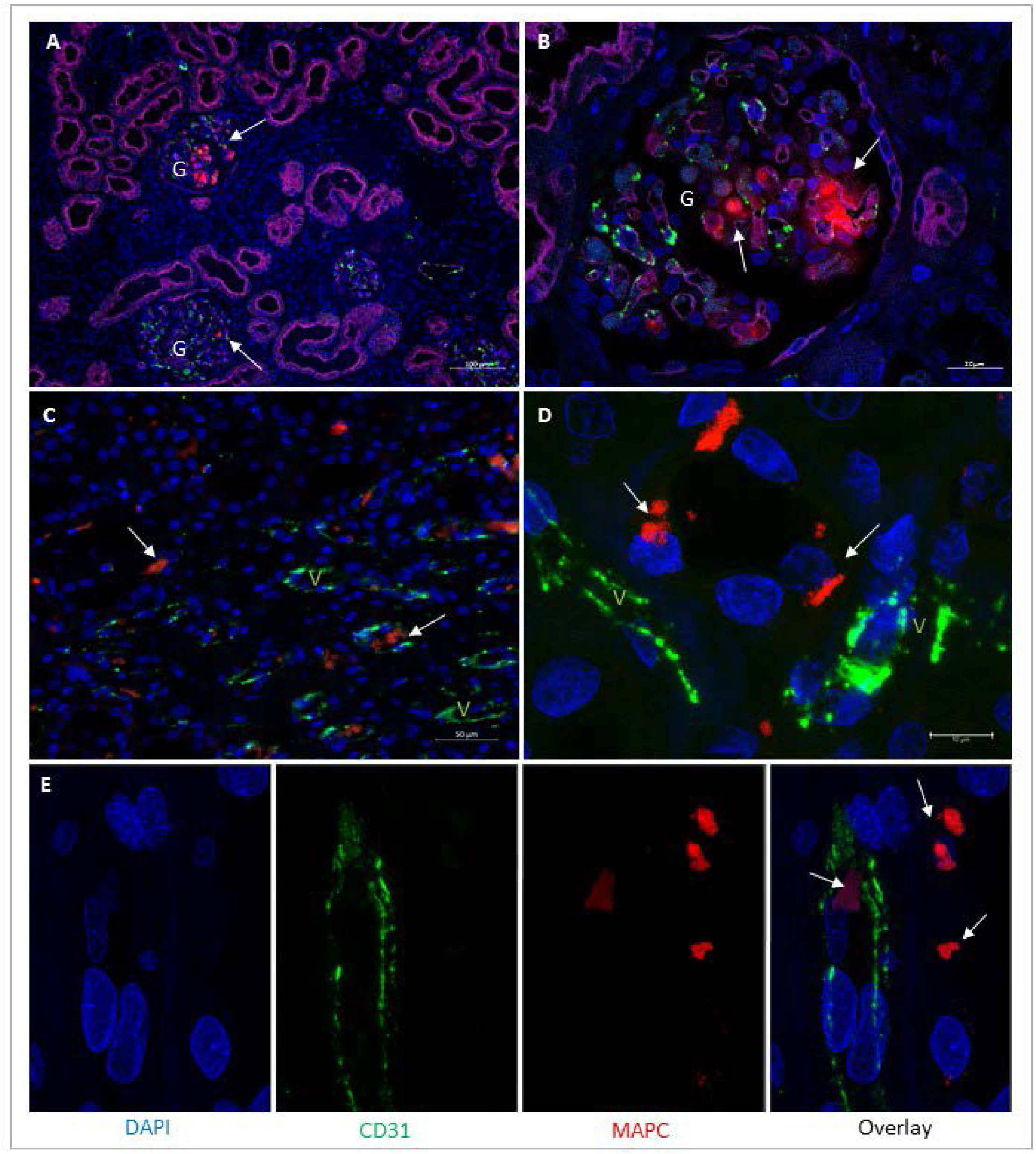
Determining MAPC cell fate and physical distribution following delivery during kidney NMP. MAPC cells were pre-labelled with a red cytoplasmic fluorescent dye (CellTracker Red CMPTX, ThermoFisher) to facilitate tracking of the cells through the kidney following 7 hours of perfusion. The red labelled cells are the MAPC cells as noted by white arrows. Counter nuclear staining in blue with DAPI. The green staining is endothelial marker CD31 and highlights the kidneys vasculature. The pink staining is Aquaporin 1 a marker for proximal tubular cells. Panels A & B are sections taken from the kidney cortex after 6 hours of MAPC treatment during NMP. These represent typical confocal microscopy image from kidneys treated with MAPC cells. The white letter (G) identifies the glomerulus. Panel C is a typical section taken from the kidney’s medulla, vessels identified by the CD31 green stain are identified by the green (V). Panel D is a high magnification image taken of 3 MAPC cells that are resident in the interstitium next to a peritubular capillary. Panel E depicts the confocal images of a blood vessel from the renal cortex with a MAPC cell in the lumen and cells that have mobilised out of the vessel into the nearby tissue.

The tissues sections could only account for the fate of MAPC cells within the kidney; there was also a proportion of MAPC cells that remained circulating within the perfusate. To evaluate this a cell filter was added to the perfusion circuit after 7 hours of NMP to capture circulating MAPC cells. Captured cells were flushed from the filter and analysed on flow cytometry, (supplementary Figure 1, Panel G). This revealed a small population of red positive cells alongside a separate population of unlabelled cells, assumed to be passenger leucocytes mobilising out of the kidney during NMP. A DAPI live/dead stain was performed and revealed the mean proportion of circulating live MAPC cells to be 21%. In contrast, the passenger leucocyte population had a mean live proportion of 44%.

## Discussion

MAPC therapy has demonstrated significant promise in treating a number of clinical conditions associated with inflammation and ischaemia ^23, 24, 25, 30^. Clinical trials have demonstrated MAPC cells have a robust safety profile. Therefore, the translation of MAPC therapy to minimise ischaemia reperfusion injury in kidney transplantation represents a promising avenue for further exploration. Here we have described the first reported series of human kidneys successfully treated with a cellular therapy using normothermic machine perfusion.

Kidneys treated with MAPC therapy demonstrated an improvement in functional parameters and biomarkers of kidney injury. A higher volume of urine output and lower urinary NGAL during NMP has previously been correlated with improved clinical outcomes following transplantation ^31, 32^. Improving these key parameters with MAPC treatment is suggestive of less tissue damage and improved cellular metabolism. We have also demonstrated a significant improvement in microvascular perfusion during NMP using CEUS. The improved blood flow seen in MAPC treated kidneys could be mediated by PGE2 or IDO, both potent vasodilators, which are reported to be part of the MAPC cell secretome in pro-inflammatory environments ^33^.

Cytokine profiling revealed that MAPC therapy during kidney NMP was associated with an anti-inflammatory, pro-tolerogenic cytokine profile. This included decreased expression of IL-1β, a pro-inflammatory cytokine associated with endothelial activation. Our group have previously correlated lower levels of IL-1β during ex vivo lung perfusion with better clinical outcomes following lung transplantation ^34^. There was also upregulation of pro-tolerogenic, anti-inflammatory IL-10 and this was accompanied by increased IDO activity. MAPC cells modulating differential cytokine expression in perfusate may be key in their potential role of minimising IRI.

Tracking the MAPC cells after delivery in the NMP circuit has revealed cells could be found throughout the kidney. Previous studies demonstrated MAPC cells have a powerful ability to home to hypoxic, damaged areas of the endothelium where they migrate into the tissue, taking up residence within the perivascular space ^35, 36, 37^. Indeed, in a pre-clinical study of acute myocardial infarction the cells actively migrated into the ischaemic, damaged myocardium following transarterial delivery ^38^. This may be mirrored in our NMP model - MAPC cells are homing to hypoxic areas within a marginal kidney where they take temporary residence. In this environment, the cells release an anti-inflammatory secretome which is having beneficial effects, resulting in increased blood flow, improved urine output and reduced tubular injury. The proportion of circulating non-viable MAPC cells may also have an important immunomodulatory role. Previous studies have demonstrated that apoptotic MSCs induce a significant immunosuppressive response mediated by recipient phagocytes ^39^.

The findings of this study reflect similar work investigating pig kidneys treated with human mesenchymal stromal cells (MSC) during NMP ^40, 41^. These studies have similarly demonstrated tracked cells mainly localising to the glomeruli. However, the authors are yet to demonstrate the reconditioning capability and changes to the inflammatory milieu that we have described in this study. This may be due to the limitations of the xenogeneic experimental model.

NMP provides a platform for direct delivery of MAPC therapy, overcoming the limitations of systemic delivery. Systemic intravenous delivery often results in entrapment of cells in the lungs or liver of the recipient, reducing delivery to the target site ^42, 43^. During NMP, the pro-inflammatory marginal kidney provides a microenvironment to prime MAPC cells, licensing them to take on the desired immunosuppressive phenotype. The combination of a direct delivery platform that can correctly license cells could provide the solution to a previous significant barrier in MSC/MAPC therapy translation ^44^.

However, there are a number of limitations in this study. NMP as an experimental system is limited in its ability to measure the post-transplantation effects of a therapy and these issues would need to be addressed in further studies. Additionally, we cannot determine the fate and persistence of the MAPC population after transplantation of the kidney. Other animal studies investigating systemic MSC therapy have demonstrated cells are no longer traceable in the recipient at 21 days, indeed the majority are lost from the system by 72 hours ^30^. Also, determining the immunogenicity of the MAPC cells in the NMP setting would be important. It has been previously demonstrated that the *in vitro* immunosuppressive activity of MAPC cells was independent of MHC matching and therefore they could be used as a universal donor product ^45^. In other clinical circumstances where allogeneic MAPC cells have been used there was no evidence of development of new anti-HLA antibodies in 16 previously unsensitised patients treated after myocardial infarction ^24^ or in 18 immunosuppressed bone marrow transplantation patients ^22^. These studies are promising, however, understanding if these positive findings would be replicated with NMP delivery will be essential going forward.

In 2018-19, 320 kidneys were not transplanted in the United Kingdom. The vast majority of these organs were rejected because of anticipated failure at reperfusion from excessive IRI. In addition, an unknown number of kidneys were not retrieved because of fears over primary non-function, despite adequate function in the donor prior to death. Previous attempts at discovering a pharmacological therapy to effectively minimise IRI have been unsuccessful (46) due to the high levels of redundancy inherent in IRI pathways. We have shown that MAPC therapy combined with EVNP interacts with multiple pathways and improves physiological function in a measurable and reassuring way. Based on these results we believe up to 50% of discarded kidneys and an unknown number of un-retrieved kidneys could be transplantable with MAPC therapy during EVNP

## Conclusion

This is the first reported series of cell therapy successfully delivered directly to human donor kidneys in an isolated *ex vivo* perfusion platform. Kidneys treated with MAPC cells during NMP demonstrate improvement in clinically relevant functional parameters and a reduction in injury and pro-inflammatory biomarkers. This may be mediated by changes to circulating cytokines or through secreted soluble anti-inflammatory mediators. NMP represents a novel cell therapy delivery system. This is a paradigm shift, providing an exciting opportunity to directly treat organs prior to transplantation to minimise ischaemia reperfusion injury. A future clinical trial evaluating this modality of delivery could result in the transplantation of otherwise discarded organs, thereby reducing the transplant waiting list and offering hope to patients with renal failure.

## Materials and Methods

### Ethics

Ethical approval for the use of human donor kidneys declined for transplantation was granted by local Research Ethics Committee (16/NE/0230). Consent for recruitment of the organ into a research study was obtained from the donor’s next of kin by specialist nurses in organ donation. Currently 15% of kidneys retrieved from organ donors across the UK are deemed unsuitable for transplant and can be offered for research purposes. The allocation of a kidney to this research project was overseen by NHS Blood & Transplant’s Research Innovation and Novel Technologies Advisory Group (RINTAG).

### Human Kidney Normothermic Machine Perfusion

On arrival at our centre kidneys were surgically prepared on ice. The renal artery was cannulated to facilitate connection to the NMP circuit. The ureter was also cannulated so urine output could be measured and sampled. Kidney pairs were perfused simultaneous for 7 hours with an oxygenated red-cell-based perfusate at a mean temperature of 36.5°C and mean arterial pressure of 75mmHg, according to published protocols ^46^. The volume of perfusate in the circuit was kept constant by matching the urine output with a crystalloid solution via continuous infusion. All physiological parameters accessible during NMP were recorded and analysed including: perfusate blood gas analysis, biochemical analysis, urine production, flow rate, and scored according to the validated quality assessment tool (7).

### MAPC treatment

The MAPC cells used in this study were obtained in collaboration with Athersys Inc, Ohio, and were provided as pre-dosed cryovial aliquots of 25×10^6^ cells. The MAPC cells were research grade Human MultiStem® cultures that were isolated from human bone marrow with consent from a healthy donor as previously described ^47^. Prior to release, the MAPC cells were confirmed to express phenotypic markers ^48^, be of >95% purity, and were fluorescently labelled with cytoplasmic dye CellTracker Red CMPTX (ThermoFisher).

For MAPC treatment of kidneys during NMP, following 60 minutes of perfusion, the right or left kidney was randomly allocated to a prescribed MAPC dose (50×10^6^ cells). In the 60 minutes preceding infusion the MAPC aliquots were gently thawed in a water bath at 37°C and then kept on ice until ready for delivery. Immediately prior to delivery cells were re-suspended in 10ml of perfusate. Control kidneys simply received a 10mL bolus of perfusate. The chosen MAPC dose was extrapolated from previous phase 2/3 clinical trials using MAPC cells for systemic infusion and calculated based on the average weight of a human kidney and volume of fluid in the perfusion circuit ^27^.

### Contrast Enhanced Ultrasound

Contrast Enhanced Ultrasound (CEUS) was performed during NMP. This technique utilises microbubbles of inert gas (sulphur hexafluoride in a phospholipid shell) to increase ultrasound signal return. Each bubble is approximately 2-3μm in diameter, allowing it to pass through the capillary bed but not the interstitium. CEUS has made it possible to assess the distribution of perfusion at a microcirculation level^49^. We have previously had success with this technique on porcine kidneys on a cold perfusion circuit and in NMP livers (16). This technique investigated whether MAPC cells cause microemboli within the microcirculation by comparing CEUS on kidneys pre and post MAPC infusion. One minute CEUS clips obtained were analysed using QLab 8.1 (Philips, Bothwell, WA, USA) CEUS quantification software. CEUS was performed using a Philips EPIQ7 Ultrasound machine. During NMP CEUS recordings were taken at 60 minutes (before MAPC infusion) and 4 hours later. A sterile sheath covered the CEUS probe and it was placed directly onto the perfusing kidney. The probe was kept stationary in a longitudinal section during contrast infusion. Contrast media (0.5mL bolus of Sonovue) was delivered via a 3-way tap on the renal artery cannula. Cineloops were acquired of the renal parenchyma and 3 standardised 5mm regions-of-interest (ROI) were captured (the interlobar artery, cortex and medulla) to record perfusion gradients. To quantify perfusion during CEUS, the Area Under the Curve (AUC) was calculated for each ROIs. A ratio of the parenchymal ROI AUC (medulla or cortex) was calculated in reference to the vessel ROI. These ratios (rAUC) were used to quantify differences in tissue perfusion. The rAUC from the start and end of perfusion were compared.

MicroFlow Imaging Ultrasound was also performed using the Philips EPIQ7 machine. MFI Ultrasound is designed to detect blood flow within the small vessels at high resolution with minimal artefact. This is displayed using overlaid blue Doppler signal and can aid in subjective interpretation of perfusion.

### ELISA

Commercially available sandwich ELISA kits (R&D) were used to measure levels of IL-8 (DY208-05) in perfusate and KIM-1 (DY1750B) & NGAL (DY1757) in the urine of the perfused kidneys according to manufacturer’s instructions.

### Mesoscale Discovery™ Multiplex

To facilitate the measurement of multiple proteins of interest in one assay, a custom electrochemi-luminescent Mesoscale Discovery™ (MSD) Multiplex was performed on perfusate samples to quantify cytokine expression during kidney perfusion. The 10-plex panel includes: IL-1β, IL-1α, IL-2, IL-6, IL-10, IL-17, TNF-α, MIP-1, IFNγ & IL-8. These assays were performed according to manufacturer’s instructions alongside technical support from an experienced MSD technician. The method of detection utilised an electrochemi-luminescent label conjugated to the detection antibody.

### High Performance Liquid Chromatography

Culture supernatants and perfusate were acidified with sodium acetate, pH=4, to give a final concentration of 15mM. Protein was precipitated with 1:10 perchloric acid 60%; samples were centrifuged, filtered and, then, analysed by HPLC to quantify kynurenine and tryptophan as described ^50^.

### In vitro endothelial cell line perfusate stimulation model

An Immortalised endothelial cell line (HMEC-1 cells, ATCC) was used to investigate the impact of the MAPC perfusate secretome on endothelial function. HMEC-1 cells were cultured to confluence and were treated with acellular perfusate diluted in modified MCDB131 media. In chamber slides, cells were stimulated with 25µl of acellular perfusate to evaluate protein expression using immunohistochemistry. In 6-well plates, HMEC-1 cells were stimulated with 250 µl of acellular perfusate to evaluate gene expression using RT-qPCR. In both situations cells were stimulated for 4 hours. RNA extraction was performed on-column using Qiagen RNeasy Plus Mini-kit as per manufacturer’s instructions. cDNA synthesis was carried out using the Tetro cDNA synthesis kit. RNA sequence quantification was carried out using TaqMan RT-qPCR on a StepOnePlus™ Real-Time PCR System. Per well 2μl of cDNA, 7μl RNase free water, 10μl SensiFAST™ Probe Hi-ROX Kit and 1μl of primer (ICAM1 TaqMan Hs00164932_m1, S1PR1TaqMan Hs00173499_m1) was added in a 96 well PCR plate. GAPDH was utilised as a housekeeping gene (TaqMan Hs02786624_g1).

### Mouse Intraperitoneal Chemotaxis Assay

All animal experiments were performed in accordance with the Home Office regulations (PPL60/4521). Eight-week-old female BALB/c mice received an intraperitoneal injection of 0.5mL of crystalloid (Ringer’s lactate), control or MAPC perfusate (n=5 mice per group). An additional 5 mice were not injected. Mice were euthanized after 6 hours treatment and 8ml of PBS-EDTA was injected into peritoneal cavity for lavage. A midline incision was made and lavage fluid collected. Lavage samples were centrifuged at 500g for 5 mins, the supernatant discarded and re-suspended in 100ul of FACS buffer ready for staining. For characterisation of leucocytes the cell suspension was stained with Ly6g-APC (Biolegend 127613) for neutrophils and CD45-PE (Biolegend 304007) to identify all leucocytes. This was incubated at 4°C for 1 hour. The cells were then washed in 200ul of FACS buffer, spun down and re-suspended in 100ul. Analysis was performed on Accuri Flow Cytometer at a rate of 14ul/min, recording 1000000 events. Results were analysed using Flow Jo software.

### MAPC tracking in kidney tissue

MAPC cells were pre-labelled with CellTracker CMPTX Red fluorescent dye (CT24552) facilitating cell tracking to evaluate partitioning and engraftment within the organ fluorescence microscopy. Core biopsies were taken at time zero, one hour, 4 hours after cell infusion and wedge biopsies at the end of perfusion. Wedge biopsies were taken by bisecting the kidney and large sample segments of cortex, medulla, collecting system and vessels. Samples were fixed in formalin and stored as paraffin embedded blocks. These were cut into 4µM sections at a later date for fluorescence microscopy imaging using a Zeiss Axioimager.

### Immunofluorescence co-localisation staining of tissue sections

Cut sections of perfused kidneys were de-waxed for 10 minutes in Xylene. The sections were then rehydrated through graded ethanol (99%, 90%, and 70%). Sections were placed in a container into a pressure cooker with 1.5L of Tris/EDTA and heated for 2mins. Blocking was performed with 10% Goat serum, 100ul per slide for 30 minutes at room temperature. Blocking buffer was removed and 100ul of primary antibody diluted in added ((S1PR1, ThermoFisher Scientific PA1-1040, 1:50; CD31, Abcam, ab182981, 1:2000; Aquaporin-1 Abcam ab168387, 1:100). Slides were incubated in the fridge at 4°C overnight. The primary antibody was washed three times in 0.05% Tween for 5 minutes. Then 100ul of the relevant secondary antibody (anti-rabbit Cy5 ThermoFisherer Scientific 1:100; anti-rabbit FITC, Abcam, ab97050 1:200, anti-rabbit Dylight550, Immunoreagents 1:100) was added and incubated for 1hour at room temperature. Following a further 3 washes, slides were incubated with 0.1% Sudan Black B in 70% EtOH to quench tissue auto-fluorescence. Coverslips were mounted with Vectashield Antifade Mounting medium with DAPI (Vectorlabs) and sealed. Since fluorescent staining fades over time it was ensured that sections were imaged and analysed taken within 2 weeks. Fluorescent imaging was performed using a Zeiss Axioimager or for high resolution Leica SP2 UV AOBS. Images were processed and analysed using Zen, LAS X or Fiji software.

### Cell filter studies

At the end of the NMP experiment a leucocyte filter (Haemonetics™) was attached to the perfusion circuit in line with the direction of blood flow. This was kept in place for 15 minutes and then disconnected. To washout the erythrocytes 250ml of cold PBS was flushed through the filter in the same direction as the blood flow. To capture the leucocytes and MAPC cells 250ml of cold PBS was then flushed in a retrograde direction and the fluid collected. This fluid was spun down in a 4°C centrifuge at 500G for 10min. To quantify the cell population the pellet was resuspended in 150ul of FACS buffer. For live/dead stain the samples were incubated with 1:2000 DAPI for 15 minutes prior to analysis. Flow cytometry was performed using the BD Fortessa X20 and the results analysed using FlowJo™ software.

### Statistical analysis

Continuous variables are reported as mean ± standard deviation where appropriate. When comparing between control and MAPC treated kidney pairs from the same donor a two-sided paired t-test was used. If another treatment group was included analysis was carried out using one-way analysis of variance (ANOVA) with a Dunnett’s post-hoc multiple comparisons correction for continuous data. When recording multiple variables over time from the same kidney a repeated measures two-way ANOVA with appropriate matching was used with Sidak’s post-hoc test for multiple comparisons. P-values of less than 0.05 were deemed statistically significant. Analyses were performed using GraphPad Prism 8.0.

## Data Availability

All data related to this study are present in the paper or the Supplementary Materials. All reagents and perfusion consumables were commercially available. MAPC cells were obtained through the material transfer agreement between Newcastle University and Athersys Inc

## Acknowledgements

We thank all the member of the perfusion and theatre team at the Freeman Hospital for technical support.

## Funding

This study was supported by Kidney Research UK, the National Institute for Health Research (NIHR) Newcastle Biomedical Research Centre and the NIHR Blood and Transplant Research Unit in Organ Donation and Transplantation at the University of Cambridge, in collaboration with Newcastle University and in partnership with National Health Service Blood and Transplant (NHSBT). The views expressed are those of the authors and not necessarily those of the National Health Services.

## Author contributions

ET, AF, NS, SA & CW conceived the study. ET, LB, IKI, TG, GW, EAI, SJT, RF, BS, AM, KC & HdL performed the experiments and data analysis. AS, SA & MLN provided technical expertise and training in perfusion. VDR & AT provided the MAPC cells and scientific advice during the project. ET, NS, SA & CW prepared and revised the manuscript. All authors read and approved the final manuscript.

## Competing interests

VDR and AT are employees of ReGenesys and Athersys, Inc, respectively and are both shareholders of Athersys stock. The other authors declare that they have no competing interests.

## Supplementary Material

**Supplementary figure 1:**
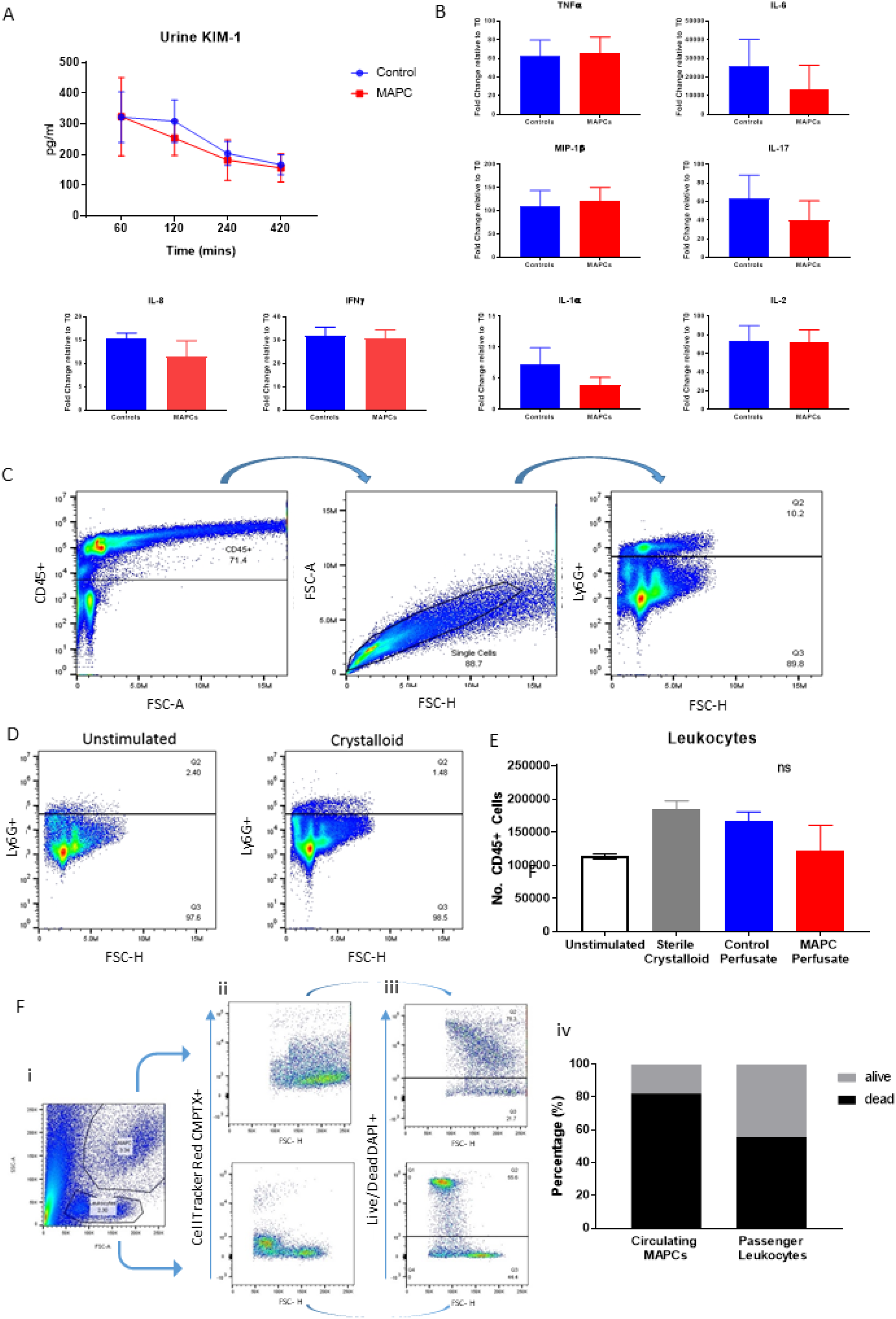
Panel A depicts urinary biomarkers of renal injury (KIM-1) measured serially during the perfusion timeline. Panel B depicts impact of MAPC therapy on the other cytokines included in the MSD panel (TNFa, IL-6, IL-1α, MIP-1β, IL-17, IL-2, IL-8 & IFNƴ). Panel C depicts the gating strategy for the mouse intraperitoneal chemotaxis experiments. Panel D depicts the corresponding flow plots for Ly6G+ cells in the unstimulated and crystalloid control groups from the mouse intraperitoneal chemotaxis experiments. Panel E is a graph demonstrating the mean number of CD45+ cells isolated from the peritoneal lavage samples following 6hour stimulations. Panel G (i-iv) depicts the results from the cell filter flow cytometry experiments designed to isolate circulating MAPCs from the perfusate at the end of NMP. Panel G (i) is the flow cytometry scatter plot of forward vs side scatter and demonstrates two distinct cell populations. Panel G (ii) examines these two cell populations staining for CellTracker Red CMPTX the MAPC fluorescent label. Panel G (iii) demonstrates a DAPI live/dead stain to quantify the proportion of live circulating cells. Panel G (iv) depicts the mean proportion of circulating cells within these 4 these four categories.

## Notes

### Author Declarations

All relevant ethical guidelines have been followed and any necessary IRB and/or ethics committee approvals have been obtained.

Any clinical trials involved have been registered with an ICMJE-approved registry such as ClinicalTrials.gov and the trial ID is included in the manuscript.

